# Behavioral and psychological symptoms of dementia: insights from a multivariate and network-based brain proteome-wide study

**DOI:** 10.64898/2026.04.23.26351110

**Authors:** Selina M. Vattathil, Duc M. Duong, Marla Gearing, Nicholas T. Seyfried, Robert S. Wilson, David A. Bennett, Randall L. Woltjer, Thomas S. Wingo, Aliza P. Wingo

**Affiliations:** Department of Neurology, University of California, Davis School of Medicine, Sacramento, CA; Department of Biochemistry, Emory University School of Medicine, Atlanta, GA; Center for Neurodegenerative Disease, Emory University School of Medicine, Atlanta, GA; Department of Pathology and Laboratory Medicine, Emory University, Atlanta, GA; Goizueta Alzheimer’s Disease Research Center, Emory University School of Medicine, Atlanta, GA; Department of Neurology, Emory University School of Medicine, Atlanta, GA; Rush Alzheimer’s Disease Center, Rush University Medical Center, Chicago, IL; Department of Pathology, Oregon Health and Science University, Portland, OR; Alzheimer’s Disease Research Center, University of California, Davis, Sacramento, CA; VA Northern California Healthcare System, Mather, CA; Department of Psychiatry, University of California, Davis, Sacramento, CA

**Keywords:** behavioral and psychological symptoms of dementia, Neuropsychiatric symptoms of dementia, BPSD, NPS, anxiety, protein differential expression

## Abstract

Behavioral and psychological symptoms of dementia (BPSD) are common, profoundly troubling to patients and caregivers, and difficult to treat, yet their molecular underpinnings remain poorly understood. Here, we generated a large brain proteomic dataset with nine BPSD domains assessed in life from 376 donors from three cohorts. Protein associations with BPSD were examined using complementary approaches — domain-specific BPSD, multi-domain BPSD, and latent factor modeling — and integrated via cross-cohort meta-analysis. Four proteins (NMT1, DCAKD, DNPH1, and HIBADH) were associated with anxiety in dementia and five proteins (ABL1, SAP18, PLXND1, CTRB2, and LDHD) with multi-domain BPSD or BPSD latent factors after adjusting for sex, age, and other covariates (FDR < 0.05). Additionally, eight protein co-expression networks were associated with BPSD across cohorts. Together, these results link BPSD to dysregulation of synaptic signaling, protein folding, and humoral immune response, providing a molecular framework for therapeutic discovery.

## Introduction

Alzheimer’s and related dementias are major public health concerns that have variable presentation across individuals. While dementia is formally defined by progressive cognitive decline and functional impairment, many people with dementia experience one or more behavioral and psychological symptoms of dementia (BPSD), such as anxiety, agitation, irritability, depression, apathy, psychosis, and sleep disturbance. These symptoms are extremely common, with clinically significant BPSD observed in 75% of people with dementia^1^.

BPSD are troubling to both patients and caregivers and are associated with higher caregiver burden and earlier placement in assisted living facilities^2,3^. While behavioral interventions are the first-line therapy, moderate to severe BPSD are treated with off-label use of medications that have limited efficacy but substantial risk of adverse side effects, including falls and FDA black box warnings of higher mortality^4-8^.Thus, safe and effective treatments for BPSD are urgently needed.

Recently, transcriptomic, proteomic and integrative analyses have shown that gene expression in the brain is altered in neurodegenerative and psychiatric conditions^9-14^. Genes identified by such analyses suggest relevant molecular processes and underlying mechanisms, and in turn offer potential targets for drug development. Similarly, identifying biological risk factors of BPSD can guide development of effective treatments. One study has investigated brain molecular alterations in BPSD, at the transcript level^15^, however BPSD mechanisms remain unclear.

Here, we aimed to identify proteins and molecular pathways associated with manifestation of BPSD using brain proteomes from the dorsolateral prefrontal cortex (dPFC) of 376 donors from three cohorts. Because BPSD comprise multiple symptom domains, we examined protein-BPSD associations at both the individual symptom level and multi-domain level using multivariate and latent factor analysis (**Figure 1**). To account for protein co-expression patterns, we performed proteome-wide differential expression analysis both at the single-protein and co-expression protein network levels (**Figure 1**). To accommodate demographic differences across the three cohorts, we analyzed each cohort separately and then used meta-analysis to identify shared associations.

**Figure 1.**
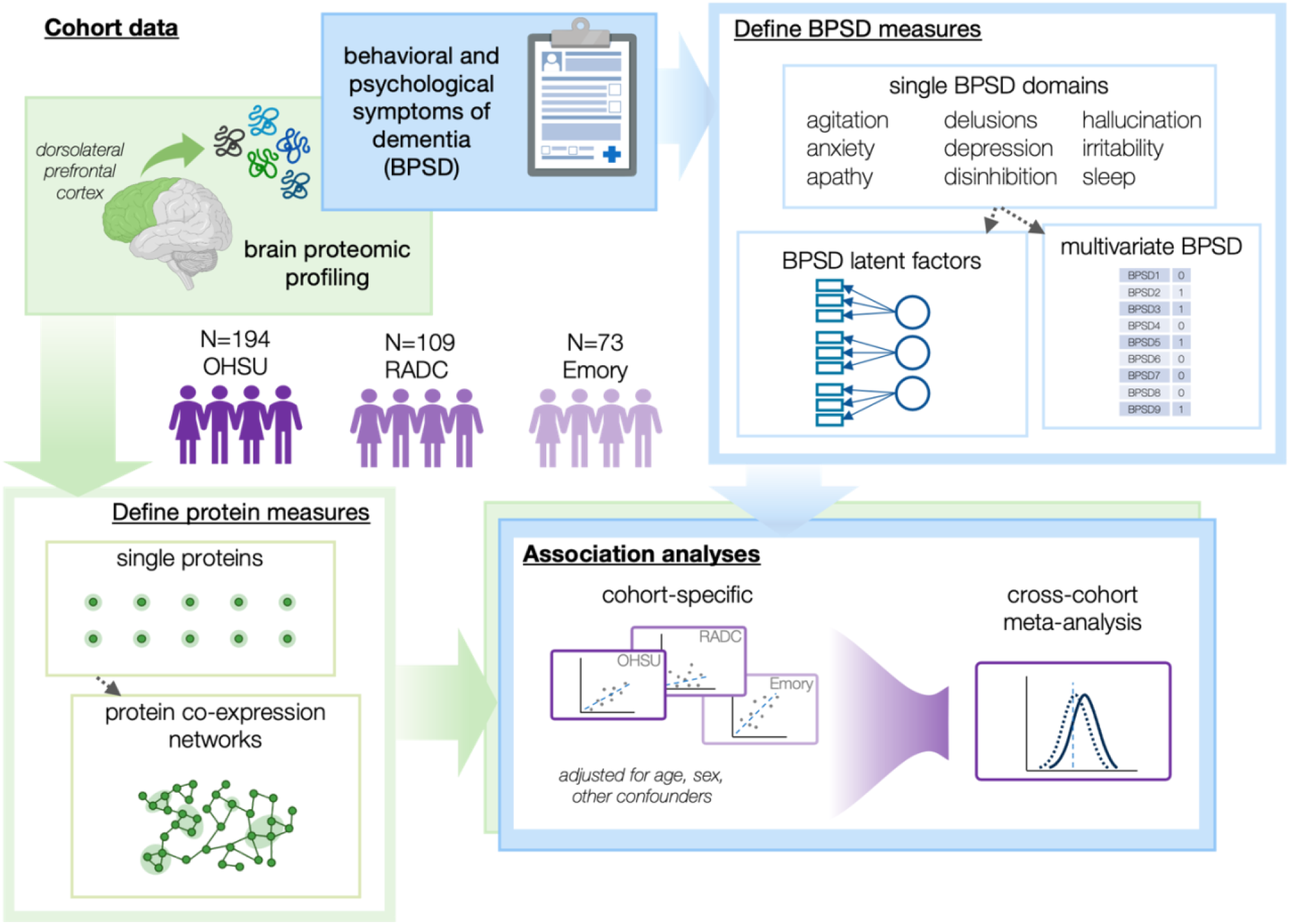
Overview of analysis. Dorsolateral prefrontal cortex samples and longitudinal measures of biological and psychological symptoms of dementia (BPSD) were collected from 376 individuals from three brain bank cohorts in the United States. Proteomic profiles and BPSD measures were analyzed to discover individual proteins and co-expression networks associated with BPSD.

We discovered four proteins associated specifically with anxiety in dementia, with consistent evidence across cohorts, five proteins linked to composite BPSD symptoms, and 8 protein co-expression modules robustly associated with one or more BPSD domains. The associated proteins suggest the role of synapse transmission, protein folding, and immune response. Collectively, these findings strengthen and expand our understanding of the molecular processes altered in BPSD and point to molecular targets for continuing research toward effective therapeutics.

## Methods

### Subjects

The study participants were from three independent research sites in the United States: Oregon Health & Science University (OHSU), Rush Alzheimer’s Disease Center (RADC) memory clinic, and Goizueta Alzheimer’s Disease Research Center at Emory University. Inclusion criteria were the presence of cognitive impairment (either mild cognitive impairment or dementia), assessment of neuropsychiatric symptoms, and available fresh frozen tissue suitable for mass spectrometry-based proteomics. To minimize confounding due to race, we grouped participants by self-reported race. Analysis was performed in only the self-reported White group due to limited sample availability in the other groups. All three studies were approved by their respective Institutional Review Boards. Consents were obtained from the participant or their legal representative.

### Behavioral and psychological symptom (BPSD) assessment

#### OHSU and Emory cohorts

BPSD were assessed longitudinally with either the Neuropsychiatric Inventory (NPI) or its short-form questionnaire (NPI-Q). We focused on the following nine symptom domains: agitation, anxiety, apathy, delusion, depression, disinhibition, hallucination, irritability, and sleep disturbance. For each domain, a participant was classified as a case if the symptom was present in the month preceding their last assessment; otherwise, the participant was considered a control.

#### RADC cohort

BPSD were assessed longitudinally using the Rush Patient Behavior Checklist and informant-based adaptation of the Hamilton Rating Scale for Depression^16,17^. From these, we extracted a subset of items that corresponded to the nine NPI/NPI-Q domains listed above and that exhibited adequate completeness in our dataset. The mapping of RADC items to each neuropsychiatric domain is presented in **Supplementary Table 1**. Each item was binary (yes=1, no=0). For a given domain, a composite score was calculated as the sum of the constituent item scores. Binary classification of the domain was then defined as follows: generally, a composite score > 0 indicated presence of the symptom (case), while score = 0 indicated absence (control). Because the composite scores for apathy were higher than for the other domains, we set the threshold at the 30^th^ percentile of the composite score distribution, and scores above this threshold were treated as cases.

### Proteomic profiling

391 participants were selected for proteomic profiling. Samples were processed in 23 batches of 17 samples each (12 batches for OHSU, 6 batches for RADC, 4 batches for Emory, plus one batch with 10 Emory and 7 RADC samples). Batches were randomized for sex, age at death, and post-mortem interval (PMI).

#### Tissue homogenization and protein digestion

Samples were homogenized in 8 M urea lysis buffer (8 M urea, 10 mM Tris, 100 mM NaH2PO4, pH 8.5) with HALT protease and phosphatase inhibitor cocktail (ThermoFisher) using a Bullet Blender (NextAdvance).The lysates were sonicated for 2 cycles consisting of 5 s of active sonication at 30% amplitude, followed by 15 s on ice. Samples were then centrifuged for 10 min at 8,000 rpm and the supernatant transferred to a new tube. Protein concentration was determined by bicinchoninic acid (BCA) assay (Pierce). For protein digestion, 200 μg of each sample was aliquoted and volumes normalized with additional lysis buffer.Samples were reduced with 5 mM dithiothreitol (DTT) at room temperature for 30 min, followed by 10 mM iodoacetamide (IAA) alkylation in the dark for another 30 min. Lysyl endopeptidase (Wako) at 1:25 (w/w) was added, and digestion allowed to proceed overnight. Samples were then 7-fold diluted with 50 mM ammonium bicarbonate. Trypsin (Promega) was then added at 1:25 (w/w) and digestion proceeded overnight. The peptide solutions were acidified to a final concentration of 1% (vol/vol) formic acid (FA) and 0.1% (vol/vol) trifluoroacetic acid (TFA) and desalted with a 10 mg HLB column (Oasis). Each HLB column was first rinsed with 1 mL of methanol, washed with 1 mL 50% (vol/vol) acetonitrile (ACN), and equilibrated with 2×1 mL 0.1% (vol/vol) TFA. The samples were then loaded onto the column and washed with 2×1 mL 0.1% (vol/vol) TFA. Elution was performed with 2 volumes of 0.5 mL 50% (vol/vol) ACN. An aliquot of 900 ul taken out and dried while the residual was used to make a global internal standard.

#### Isobaric tandem mass tag (TMT) peptide labeling

Each sample was re-suspended in 100 mM TEAB buffer (100 μL). The TMT labeling reagents (5mg) were equilibrated to room temperature, and anhydrous ACN (200μL) was added to each reagent channel. Each channel was gently vortexed for 5 min, and then 20 μL from each TMT channel was transferred to the peptide solutions and allowed to incubate for 1 h at room temperature. The reaction was quenched with 5% (vol/vol) hydroxylamine (5 μL) (Pierce). All channels were then combined and dried by SpeedVac (Labconco) to approximately 100ul and diluted with 1 mL of 0.1% (vol/vol) TFA, then acidified to a final concentration of 1% (vol/vol) FA and 0.1% (vol/vol) TFA. Labeled peptides were desalted with a 30 mg HLB column (Oasis). Each HLB column was first rinsed with 1 mL of methanol, washed with 1 mL 50% (vol/vol) acetonitrile (ACN), and equilibrated with 2×1 mL 0.1% (vol/vol) TFA. The samples were then loaded onto the column and washed with 2×1 mL 0.1% (vol/vol) TFA. Elution was performed with 2 volumes of 0.5 mL 50% (vol/vol) ACN. The eluates were split into 300 and 700ul and dried to completeness using a SpeedVac.

#### High-pH off-line fractionation

Dried samples (30%) were re-suspended in high pH loading buffer (0.07% vol/vol NH4OH, 0.045% vol/vol FA, 2% vol/vol ACN) and loaded onto a Water’s BEH 1.7 um 2.1mm by 150mm. A Thermo Vanquish was used to carry out the fractionation. Solvent A consisted of 0.0175% (vol/vol) NH4OH, 0.01125% (vol/vol) FA, and 2% (vol/vol) ACN; solvent B consisted of 0.0175% (vol/vol) NH4OH, 0.01125% (vol/vol) FA, and 90% (vol/vol) ACN. The sample elution was performed over a 25 min gradient with a flow rate of 0.6 mL/min. A total of 96 individual equal volume fractions were collected across the gradient and dried to completeness using a SpeedVac.

#### Mass spectrometry analysis and data acquisition

All samples (∼1ug for each fraction) were loaded and eluted by Ultimate 3000 RSLCnano or Vanquish Neo (Thermo Fisher Scientific) with a 150 μM I.D. capillary column with 1.7 μM C18 CSH (Water’s) over a 26 min gradient. Mass spectrometry was performed with a high-field asymmetric waveform ion mobility spectrometry (FAIMS) Pro frontend equipped Orbitrap Eclipse (Thermo) in positive ion mode using data-dependent acquisition with 1.5 second top speed cycles for each FAIMS compensative voltage (CV). Each cycle consisted of one full MS scan followed by as many MS/MS events that could fit within the given 1.5 second cycle time limit. MS scans were collected at a resolution of 120,000 (410-1600 m/z range, 4×10^5 AGC, 50 ms maximum ion injection time, FAIMS CV of -45 and -65). Only precursors with charge states between 2+ and 6+ were selected for MS/MS. All higher energy collision-induced dissociation (HCD) MS/MS spectra were acquired at a resolution of 30,000 (0.7 m/z isolation width, 38% collision energy, 125k AGC target, 54 ms maximum ion time, TurboTMT on). Dynamic exclusion was set to exclude previously sequenced peaks for 20 seconds within a 10-ppm isolation window.

#### Database search

The raw TMT files were processed to protein abundances with the FragPipe analysis suite (version 22.0 with MSFragger version 4.1), which utilizes MSFragger and Philosopher. Raw files were searched against the canonical UniProt human protein database (downloaded February 2019 supplemented with APOE 2 and 4 and Abeta 40 and 42 isoforms: 20,404 total target entries) with the default parameters for TMTPro search: fully tryptic, 2 miscleavages and 20 ppm mass tolerances. The database search yielded data for 11,223 proteins.

### Proteomics data preparation

Proteomic data were first cleaned by converting zero intensity values to missing values; zero values occur batch-wise and indicate a protein was not measured in the batch. Proteins observed in fewer than 50 participants across the three cohorts were removed. Sample abundances were rescaled to correct for differences in protein load and then log_2_-transformed to improve normality. Batch effects were regressed out of the scaled, log_2_-transformed matrix, after which principal component (PC) analysis was applied to the proteins with complete data. Samples that were more than four standard deviations from the mean for either of the first two PCs were deemed outliers; two outlier samples were removed in this step.

Following this global preprocessing, cohort-specific steps were performed. First, samples that did not have at least one BPSD observation were excluded (10 from OHSU and 4 from Emory). Then, proteins were filtered to those measured in at least 50 samples in the cohort, resulting in 10,163 proteins for OHSU,9,369 for RADC, and 7,959 for Emory. For each cohort, a linear regression model was fitted to estimate the effects of batch, PMI, and age at death. The residuals were retained as covariate-adjusted proteomic profiles for downstream analyses.

### Statistical analysis

All analyses were performed in R (version 4.4.2) unless noted. False discovery rates were estimated using package *qvalue*.

#### Latent factor analysis

Latent factor analysis was performed in each cohort separately to allow for different structures resulting from differences in BPSD measures and age at death. The binary last visit observations served as the observed variables. The approach required complete data for all domains; to preserve sample size in the RADC analysis, the delusion and disinhibition domains were dropped. First, tetrachoric correlations among the binary BPSD were estimated with package *polycor*. Exploratory factor analysis (EFA) with oblimin rotation was then performed using package *psych*. The number of factors to model was guided by the scree plot. Factor loadings from the initial model were examined for interpretability, and when indications of over-fitting emerged (e.g., single-item factors), the model was specified with fewer factors and the EFA was rerun. The identified latent factors were then used as outcomes in protein differential expression analysis as described below.

#### Proteome-wide differential expression analysis using individual BPSD

Prior to association testing, surrogate variables (SVs) that capture latent unwanted sources of variation in the proteomic data were estimated with package *sva*^18^. The number of significant SVs varied by cohort and BPSD (range 8–21). The SVs were regressed out of the covariance-adjusted proteomic profiles described in the previous section, and the resulting residuals were taken as SV-corrected protein abundances. For each BPSD, association between BPSD and protein abundance was tested using logistic regression, with BPSD represented by binary last visit observation and self-reported sex (male v. female) included as a covariate. Analysis was performed within each cohort, then proteins measured in all three cohorts were carried forward to meta-analysis. Specifically, cohort-specific effect estimates and standard errors were combined using the fixed-effects inverse-variance method implemented in METAL^19^. Proteins with FDR < 5% were deemed significantly differentially expressed for the given BPSD.

#### Proteome-wide differential expression analysis using latent factors of BPSD

Each latent factor in each dataset was tested for association with brain protein abundance using linear regression, with sex and protein surrogate variables as covariates. Multiple testing correction was performed in each run, and significant differential expression was defined at FDR < 5%.

#### Proteome-wide differential expression analysis using multivariate BPSD outcome

Differential expression analysis using all nine binary BPSD as a multivariate outcome was performed with a linear mixed model using the geeglm function from package *geepack*, including sex as a covariate in the model. Models were fitted in each cohort, then results were meta-analyzed using package *mixmeta*. To preserve sample size in the RADC analysis, the delusion and disinhibition domains were dropped.

#### Protein co-expression modules

Protein co-expression modules were constructed using WGCNA^20^. Proteins were represented by covariate-adjusted proteomic profiles. To build a consensus network across all three cohorts, we used the blockwiseConsensusModules function to construct the consensus topological overlap matrix (TOM) as the element-wise minimum across cohort-specific TOMs. We chose a soft power threshold of 8 based on scale-free topology fit diagnostics in each cohort. The other WGCNA parameters were specified as follows: networkType=signed, TOMtype=signed, corType=bicor, pamRespectsDendro=FALSE, mergecutheight=0.3, deepSplit=4, reassignThreshold=0, maxPoutliers=0.1. The biological significance of each module was inferred by gene set enrichment using package *clusterProfiler*. In each cohort, protein modules were summarized by their eigengenes and tested for association with BPSD using regression models with sex as a covariate. Association results for the individual BPSD domains were combined across cohorts using fixed-effects meta-analysis. Significant association was defined at p-value < 0.05.

## Results

### Sample characteristics

The analysis included 376 participants from three cohorts (**Table 1)**. All participants had a clinical diagnosis of dementia at time of death, except for one with MCI. All RADC participants (N=109) had BPSD observations within two years of death. In the OHSU (N=194) and Emory (N=73) datasets, the percentage of participants with BPSD observations within two years of death was 77% and 58%, respectively.

**Table 1.** Characteristics of the participants in the OHSU, Emory, and RADC datasets.

|  | OHSU (N = 194) |  |  | RADC (N = 109) |  |  | Emory (N = 73) |  |  |
| --- | --- | --- | --- | --- | --- | --- | --- | --- | --- |
|  | N | % |  | N | % |  | N | % |  |
| Sex |  |  |  |  |  |  |  |  |  |
| male | 81 | 42% |  | 47 | 43% |  | 42 | 58% |  |
| female | 113 | 58% |  | 62 | 57% |  | 31 | 42% |  |
|  | N cases | % cases | N missing | N cases | % cases | N missing | N cases | % cases | N missing |
| agitation | 61 | 31% | 0 | 53 | 49% | 0 | 41 | 56% | 0 |
| anxiety | 70 | 36% | 0 | 74 | 68% | 0 | 29 | 40% | 0 |
| apathy | 60 | 31% | 0 | 70 | 65% | 2 | 38 | 52% | 0 |
| delusion | 19 | 10% | 0 | 28 | 34% | 27 | 12 | 16% | 0 |
| depression | 81 | 42% | 0 | 26 | 24% | 0 | 21 | 29% | 0 |
| disinhibition | 29 | 15% | 0 | 23 | 34% | 42 | 21 | 29% | 0 |
| hallucination | 21 | 11% | 0 | 57 | 53% | 2 | 18 | 25% | 0 |
| irritability | 55 | 28% | 0 | 35 | 33% | 2 | 30 | 41% | 0 |
| sleep | 73 | 38% | 0 | 52 | 49% | 3 | 23 | 32% | 0 |
|  | mean (SD) | median | range | mean (SD) | median | range | mean (SD) | median | range |
| age at death | 91.3 (10.2) | 93 | 36.8-111.7 | 79.4 (8.0) | 79.5 | 60.9-96.6 | 78.2 (8.3) | 78 | 60.0-95.0 |
| PMI | 15.6 (17.3) | 11.5 | 2.0-108.0 | 6.1 (3.7) | 5 | 2.0-28.1 | 10.4 (9.1) | 7 | 2.5-64.0 |

Neuropathologic diagnosis was also available for all individuals (**Supplementary Table 2**). Overall, the prevailing dementia etiology was Alzheimer’s disease. Specifically, all Emory participants, 86% of RADC participants, and 78% of OHSU participants had a primary neuropathologic diagnosis of Alzheimer’s disease. Age at death was significantly higher in OHSU compared to Emory (t-test p=1.1 × 10^−20^) and RADC (t-test p=3.6 × 10^−24^). PMI also varied significantly between cohorts (t-test OHSU v. Emory p = 1.8 × 10^−3^; OHSU v. RADC p=4.1 × 10^−12^; RADC v. Emory p= 2.2 × 10^−4^). In all analyses, we adjusted for the effects of age at death and PMI.

Significant associations were defined at p-value < 0.05. The hub proteins for each module were defined as those with intramodular connectivity (kME) > 0.7. The top hub proteins based on kME are listed (up to 10 hub proteins per module). Enriched Gene Ontology (GO) biological processes were identified with R package *clusterProfiler*. Selected top processes for each module are listed; the full set of enriched processes is presented in Supplementary Table 6.

Case rates for individual BPSD varied across cohorts. In OHSU, depression was the most common symptom (42% of participants). In RADC, anxiety and apathy were most common (65–68%), while in Emory, agitation was predominant (56%). These differences likely reflect both variation in participant composition and the distinct instruments used to capture BPSD. To best utilize the data from the three cohorts while maintaining robust signal, we performed extensive quality control on the proteomics data from each cohort, and performed latent factor, protein co-expression, and association analyses in each cohort separately.

### Proteins associated with individual BPSD

We first examined the association between individual proteins and each of the nine BPSD domains within each cohort. Then, using the proteins measured in all three cohorts, we used fixed-effects meta-analysis to identify proteins with consistent effects across cohorts. At false discovery rate (FDR) < 5%, four proteins reached significance for anxiety – DCAKD had higher abundance while NMT1, DNPH1, and HIBADH had lower abundance in participants with anxiety relative to controls. (**Figure 2, Supplementary Table 3**).

**Figure 2.**
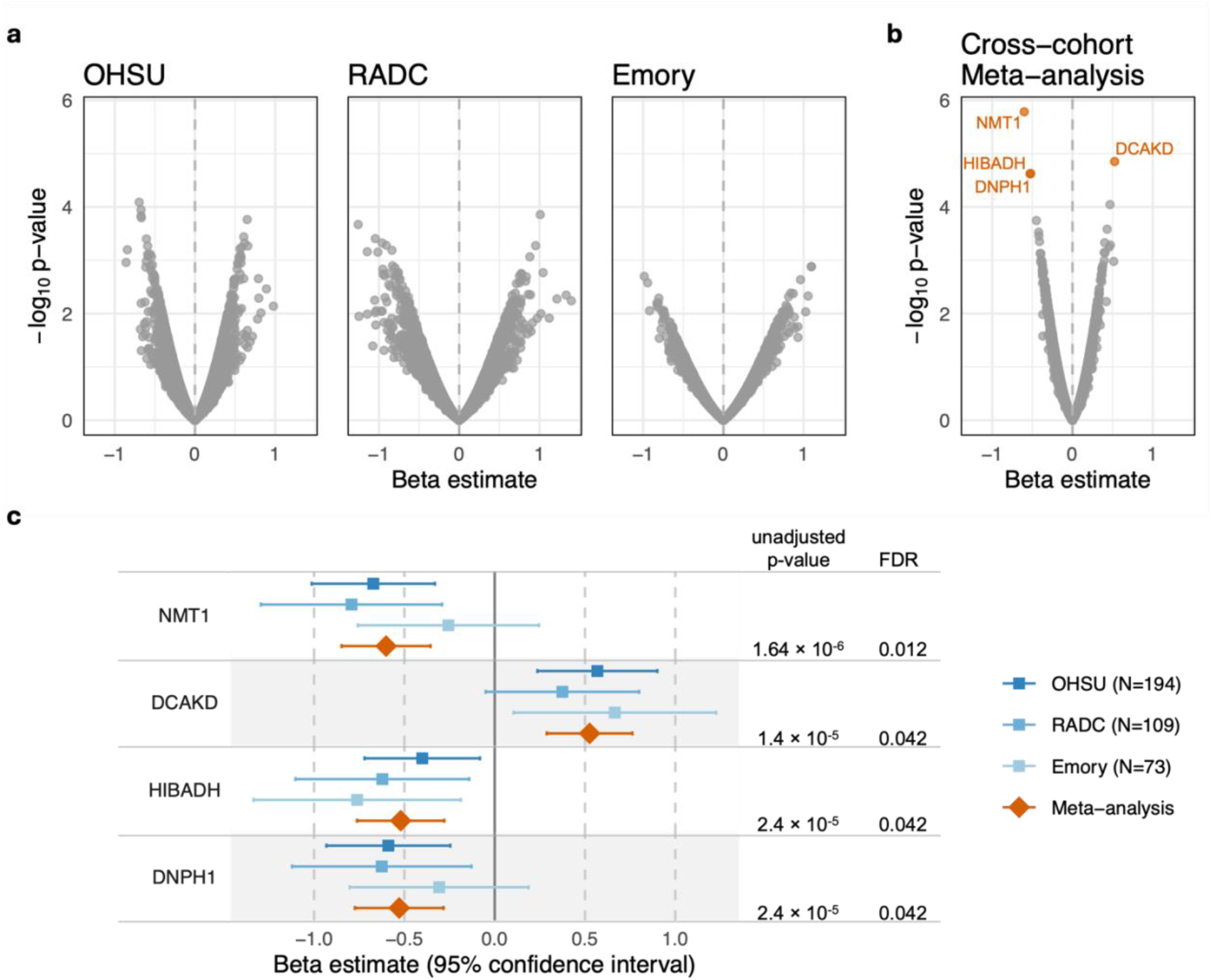
Protein differential expression results for anxiety. **a-b**, Volcano plots for (**a**) cohort-specific analyses and (**b**) cross-cohort meta-analysis. Proteins are plotted in color if they were significantly associated with anxiety in the respective dataset and plotted in gray otherwise. **c**, Forest plot of beta estimates for proteins associated with anxiety in meta-analysis. Diamond and square markers indicate point estimate of the beta coefficient, or change in log-odds associated with 1 standard deviation increase in surrogate variable-corrected protein abundance. Horizontal bars indicate 95% confidence interval of the estimate.

As a sensitivity analysis, we examined the associations after adjusting for APOE ε4 isoform expression. APOE ε4 expression itself was not significantly associated with any BPSD in our analyses and including it as a covariate made little change in the observed results for other proteins (**Supplementary Table 4**). Next, we explored potential sex-specific effects using sex-stratified analyses. No protein reached the FDR < 5% threshold for any of the nine BPSD in either males or females, indicating an absence of strong sex-specific associations in the present dataset.

### Proteins associated with latent factors of BPSD

BPSD domains are mild to moderately correlated with each other and may have shared etiology. Therefore, we used exploratory factor analysis (EFA), a data-driven approach to infer the underlying structure among observed traits, to characterize the observed BPSD patterns. We found that the most parsimonious factor structure differed across cohorts (**Figure 3a**). In the OHSU cohort, a two-factor model provided the best fit, with Factor 1 capturing behavioral and affective symptoms and Factor 2 capturing psychosis and apathy. In the Emory cohort, a three-factor model emerged, with factors corresponding to apathy, psychosis, and sleep disturbance (Factor 1), disinhibition and affective symptoms (Factor 2), and behavioral symptoms (Factor 3). In the RADC cohort, we excluded the delusion and disinhibition domains due to their higher missing data rates to preserve overall sample size. Without delusion and disinhibition, the best fit model in RADC was a single factor driven by arousal-related BPSD (agitation, irritability, anxiety), with secondary contributions from hallucination, depression, and sleep disturbance and negligible contribution from apathy.

**Figure 3.**
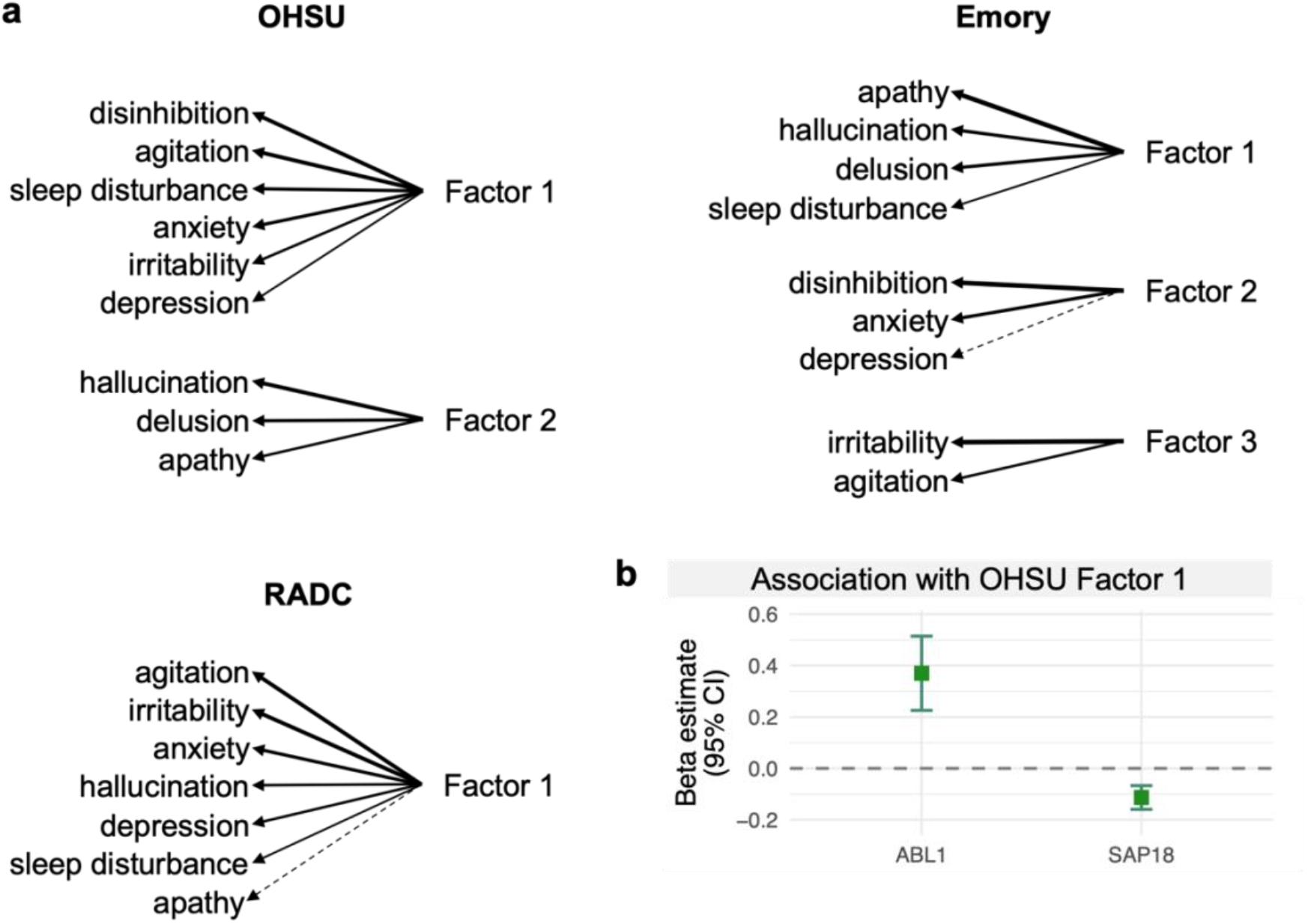
Latent factor models and differential expression. **a**. Factor loadings in OHSU, Emory, and RADC datasets. The width of the lines connecting the latent factors and the observed BPSD indicate loading strength; loadings less than 0.2 are indicated with a dashed line. Delusion and disinhibition were dropped in the RADC analysis due to high data missing rate. **b**. Effect estimates (log-odds) for the two proteins significantly associated with OHSU Factor 1.

We used the latent factor scores for each participant to identify proteins associated with the underlying factors of BPSD. Association was tested separately for each of the two OHSU latent factors, three Emory latent factors, and the RADC latent factor. We found two proteins significantly associated with OHSU Factor 1 (FDR < 5%), with ABL1 positively associated and SAP18 negatively associated with the latent factor (**Supplementary Table 5, Figure 3b**).

### Multivariate association analysis

Another approach to test protein expression with multiple BPSD domains is multivariate analysis, which considers all BPSD domains simultaneously as the phenotype of interest. Multivariate analysis tests the hypothesis that protein expression is associated with any BPSD domain, accounting for the non-independence among the domains. The cohort-specific multivariate analyses identified four proteins associated with overall BPSD manifestation—ABL1, PLXND1, and CTRB2 in OHSU, and LDHD in Emory (FDR < 5%). To understand which domains drive the association, we examined the estimated effects from the domain-specific analyses, which are visualized in **Figure 4**. For each protein, the estimated effects are uniformly positive or negative across BPSD domains. For ABL1, expression was most strongly associated with disinhibition, agitation, anxiety, and irritability. These domains were captured by OHSU Factor 1, and as expected this latent factor is also positively associated with ABL1. Like ABL1, PLXND1 was most strongly associated with disinhibition, but delusion and agitation were also among the top domains. For CTRB2, expression was most strongly associated with disinhibition, hallucination, and apathy. For LDHD, which had significant association in the Emory cohort, expression was most strongly associated with the hallucination and apathy domains, which contributed to Emory Factor 1. Overall, the findings are consistent with the inferences from the latent factor modeling.

**Figure 4.**
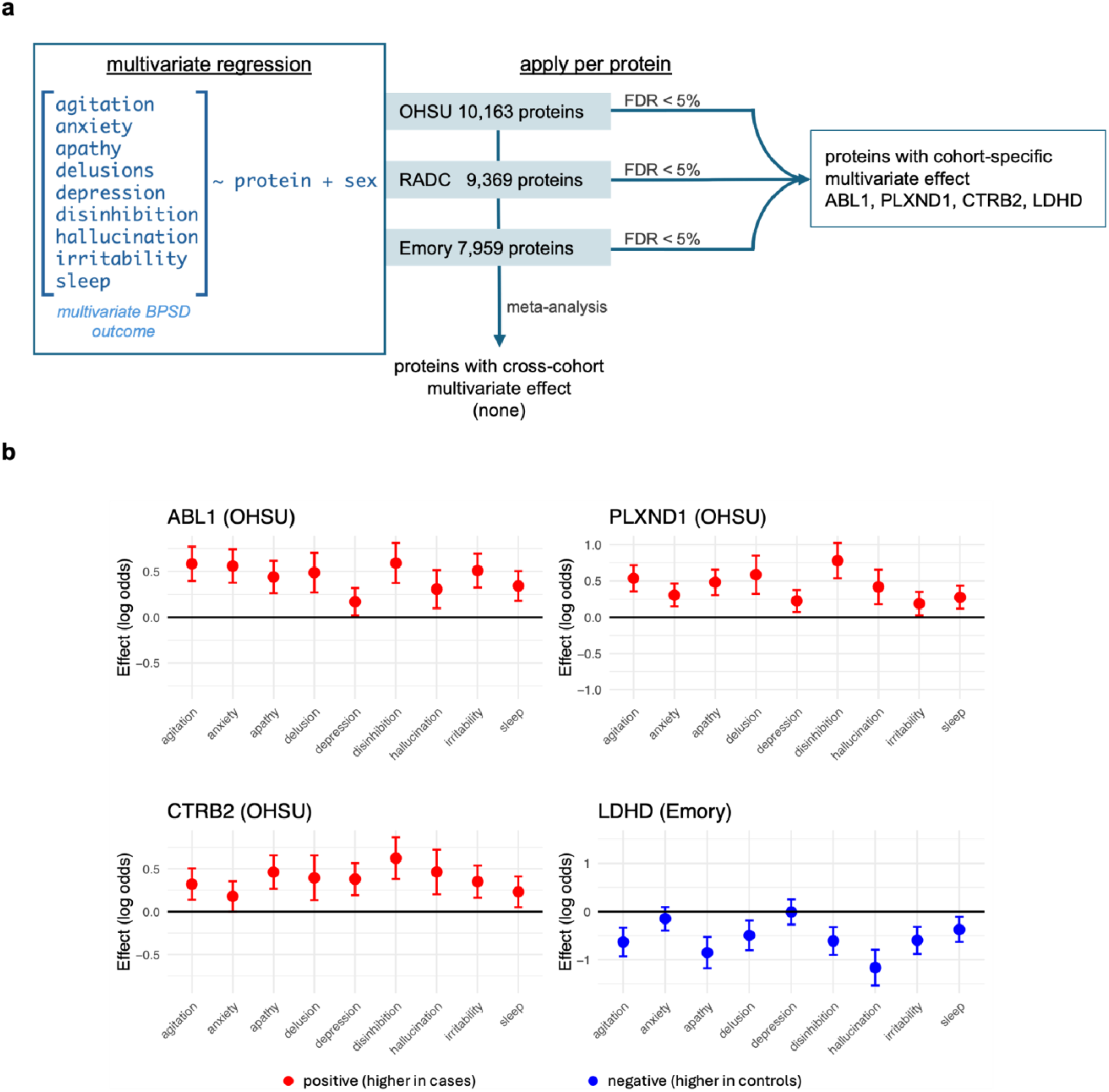
Summary of multivariate analysis. **a**. Schematic of workflow. The multivariate regression formula modeled the BPSD domains as repeated measures for each participant, and a Wald test was applied to test for protein association with the BPSD as a joint outcome. Significant proteins were identified in each cohort separately and using cross-cohort meta-analysis. **b**. Estimated effect on each BPSD domain for the four proteins with significant multivariate effect. The plots show the effect (log odds) estimates (points) and standard errors (vertical bars) in the cohort with the significant multivariate effect. For each protein, the effect estimates are uniformly positive or negative across BPSD domains.

### Protein co-expression modules associated with BPSD

Proteins operate within interconnected networks rather than in isolation; therefore, identifying co-expressed modules can reveal biologically meaningful pathways that influence phenotypic outcomes. We used weighted gene co-expression network analysis (WGCNA)^20^ to define modules of co-expressed proteins, which yielded 19 modules shared across the three cohorts **(Supplementary Tables 6-7)**. We then tested the module eigengenes for association with individual BPSD, the BPSD-based latent factors, and the multivariate BPSD outcome in each cohort **(Supplementary Tables 8-9)**. Eight modules were associated (p-value < 0.05) with one or more BPSD domains in the meta-analysis **(Table 2)**. Specifically, four modules (salmon, tan, green, pink) were associated with irritability and/or agitation and were enriched in regulation of tyrosine phosphorylation of STAT protein, humoral immune response, mitochondrial translation, and mRNA metabolic process. Three other modules (turquoise, lightgreen, magenta) were associated with depression and/or anxiety and were enriched in mRNA processing, microtubule cytoskeletal organization, and small molecule catabolic process. The grey60 module, enriched in genes involved in protein folding, was associated with apathy.

**Table 2.**
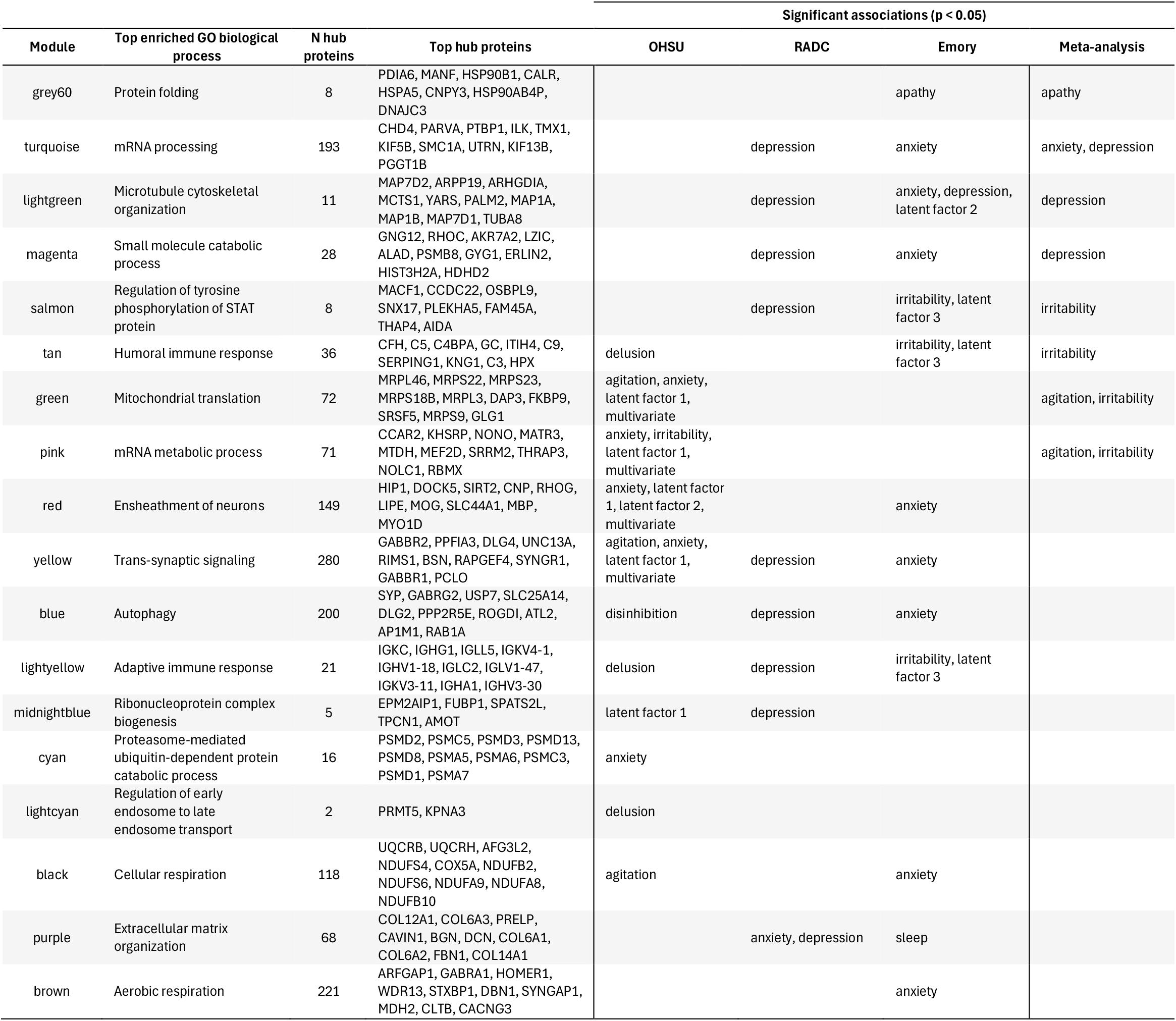
Protein co-expression modules associated with individual BPSD domains, BPSD latent factors, or multivariate BPSD. Significant associations were defined at p-value < 0.05. The hub proteins for each module were defined as those with intramodular connectivity (kME) > 0.7. The top hub proteins based on kME are listed (up to 10 hub proteins per module). Enriched Gene Ontology (GO) biological processes were identified with R package *clusterProfiler*. Selected top processes for each module are listed; the full set of enriched processes is presented in Supplementary Table 6.

While the associations from the meta-analysis are of particular interest because they have evidence that they generalize across cohorts, the associations from the cohort-specific analyses also reveal potential important biological processes important in BPSD. Some BPSD domains, particularly anxiety, were associated with multiple co-expression modules (**Figure 5**). Likewise, depression was associated with 8 protein co-expression modules, irritability with 5 modules, and agitation with 4 modules (Figure 5). With latent factors of BPSD, in OHSU, five modules were associated with Factor 1, which captured behavioral and affective symptoms. The associated modules were enriched in nervous system-related processes such as ensheathment of neurons and synaptic signaling as well as basic processes such as mRNA processing and mitochondrial translation **(Table 2)**. In Emory, modules enriched in immune response-related proteins (tan, lightyellow, salmon) were associated with the latent factor capturing behavioral reactivity (irritability and agitation). Taken together, the observations suggest some molecular processes have more robust association with BPSD while others are more subtle or context-dependent.

**Figure 5.**
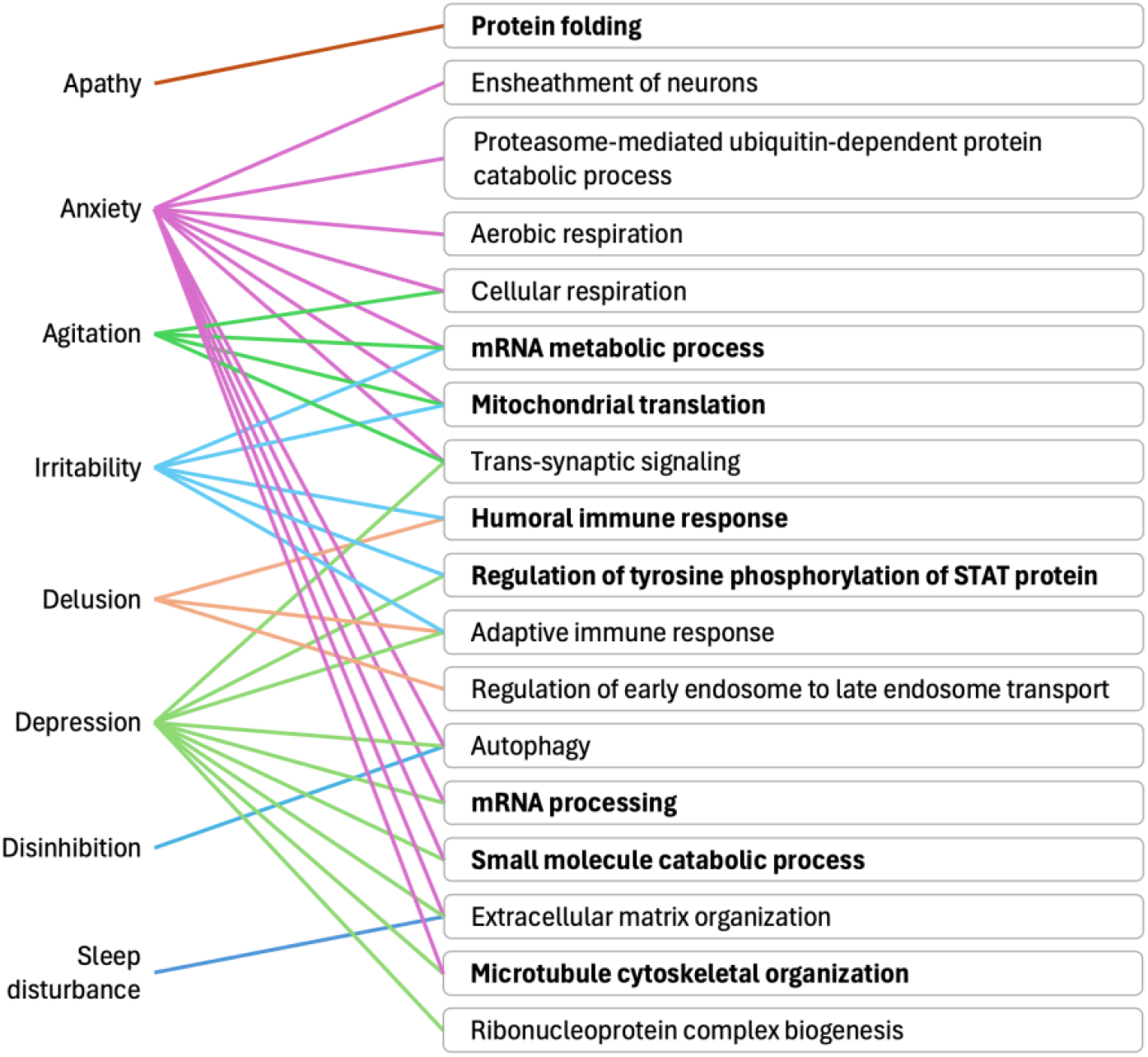
Mapping of BPSD domains to biological processes. Association between BPSD domains and protein co-expression module eigengenes was tested using linear regression. For each module, the top enriched Gene Ontology (GO) biological process was identified with R package *clusterProfiler*. Colored lines denote significant association (p-value < 0.05) between a BPSD domain and biological process in at least one cohort or cross-cohort meta-analysis. Bold text identifies biological processes associated in meta-analysis.

## Discussion

BPSD are among the most profoundly troubling aspects of living with dementia for both patients and caregivers. However, the molecular underpinnings of BPSD remain poorly understood, limiting the identification of effective therapeutic targets for these prevalent and debilitating symptoms. To address this gap, we generated human brain proteomes of 376 donors from three cohorts with nine BPSD domains assessed in life and conducted the first brain proteome-wide differential analysis of BPSD. We comprehensively examined BPSD through complementary approaches – considering each BPSD domain individually, all domains simultaneously as a multivariate outcome, and using latent factors to capture the underlying variation across domains – as well as considering proteins both individually and as co-expressed modules. Data from each cohort were analyzed separately and integrated via meta-analysis, a strategy which respected cohort-level differences while maximizing statistical power. Together, these approaches identified nine proteins (DCAKD, DNPH1, HIBADH, NMT1, ABL1, SAP18, PLXND1, CTRB2, LDHD) associated with BPSD at stringent significance thresholds (FDR <0.05) and eight co-expressed protein modules robustly associated with BPSD across three cohorts, shedding light on the molecular and biological processes that may contribute to the development of BPSD.

From the BPSD domain-specific analyses, we found that expression levels of DCAKD, NMT1, HIBADH, and DNPH1 proteins were altered in individuals with dementia who experienced anxiety symptoms. DCAKD (dephospho-CoA kinase domain-containing protein) is a membrane protein that enables dephospho-CoA kinase activity and is principally involved in coenzyme A biosynthesis^21^. NMT1 encodes a protein that catalyzes N-myristoylation, a co-translational and post-translational lipid modification that affects protein membrane targeting, stability, and function. The absence of prior literature linking DCAKD and NMT1 to BPSD or related conditions positions these proteins as unexplored candidates worthy of further investigation. On the other hand, HIBADH has been identified as a candidate causal protein in bipolar disorder and smoking behavior^22^, and genome-wide association studies have linked variants in the *HIBADH* region to suicide attempts^23^. Thus, HIBADH is likely to contribute to the pathogenesis of mood disorders, and our study further suggests that it is altered in anxiety in dementia patients. Finally, DNPH1, a 5-hydroxymethyl-dUMP N-hydrolase that removes cytotoxic nucleotides and safeguards DNA replication fidelity^24^, has been causally linked to schizophrenia^25^ and major depressive disorder^22^ through Mendelian randomization, and nearby SNPs have been associated with sleep disturbance in people with psychiatric conditions^26^. Our results add novel insight that expression of DNPH1 protein is altered in dementia patients with anxiety. In line with our findings in brain, in cerebrospinal fluid (CSF), DNPH1 was associated with anxiety, agitation, and irritability in individuals with MCI or dementia in a recent BPSD differential expression analysis of CSF proteomic profiles from 418 ADNI participants (Zhen et al, manuscript in preparation), highlighting the potential of DNPH1 as a translational biomarker.

The multivariate and latent factor analyses linked multi-domain BPSD to five proteins — SAP18 (histone deacetylase complex subunit SAP18), ABL1 (tyrosine-protein kinase ABL1), PLXND1 (plexin D1), CTRB2 (chymotrypsinogen B2), and LDHD (lactate dehydrogenase D). ABL1 is highly expressed in neurons and has been implicated in neurodegenerative disease through its effect on synaptic structure and plasticity^27^. PLXND1 has been linked to neuronal structure and function, specifically axonogenesis and synapse assembly. Consistent with these findings, we found that synaptic signaling and ensheathment of neurons were top enriched processes for protein modules associated with BPSD (agitation, anxiety, depression, and OHSU latent factor 1). Together, our protein-specific and co-expression module analyses strongly point to the role of synaptic structure and synaptic signaling in BPSD.

From the co-expression module analyses, eight additional biological processes emerged as being altered in BPSD across the three cohorts. These included mRNA processing, mRNA metabolic process, protein folding, regulation of tyrosine phosphorylation of STAT protein, microtubule cytoskeletal organization, catabolic process of small molecules, humoral immune response, and mitochondrial translation. Immune response has been implicated in psychiatric disorders^28^ and neurodegenerative diseases^29^, and now our findings link humoral immune response to BPSD. Protein folding was altered in apathy in AD, and the top hub protein in the enriched module is PDIA6, an enzyme that promotes protein folding and helps target misfolded proteins for refolding or degradation and has been observed within neurofibrillary tangles in AD brain ^30-32^. Previous work has found that tau burden was associated with longitudinal apathy burden in people with MCI and AD^33^. Taken together, the association we observe supports the idea that proteins involved in protein folding and reducing tau burden are potential targets for treating apathy in people with MCI and AD.

We recognize limitations of our study. First, our analyses used proteomic profiles from dPFC. The dPFC is frequently used for studies of Alzheimer’s disease since it is less affected by cell death in later stages of the disease than other regions, but it may not be optimal for studying BPSD. This possibility is supported by a recent transcriptome-wide differential expression analysis for post-traumatic stress disorder (PTSD) and major depressive disorder that used data from both dPFC and dorsal anterior cingulate cortex (dACC) in 325 donors^13^. While the differential expression results from dPFC and dACC were highly correlated, more genes were significantly differentially expressed in dACC than dPFC for both PTSD (16 genes v. 1 gene) and major depression (67 genes versus 1 gene)^13^. This highlights that future BPSD studies should investigate the anterior cingulate cortex and other brain regions. Second, we included only donors who self-identified as White due to low availability of donors from other groups. Analyses in other groups are needed to make conclusions that are applicable to the wider population. Third, BPSD was assessed with multiple questionnaires in the Rush cohort and with the NPI-Q in OHSU and Emory cohorts. Ideally the BPSD would be measured using standardized criteria across cohorts, which would enable investigation of inconsistencies between cohorts while also boosting power to detect shared effects. This reflects the challenges of having post-mortem brain donors with BPSD assessment in life, particularly in several domains of BPSD beyond depression. Nevertheless, we mitigated the impact of cross-cohort differences by using meta-analysis to identify protein candidates associated with BPSD across cohorts.

In summary, this brain proteomic study of nine BPSD domains links proteins and biological pathways to BPSD and provides an unprecedented resource for the molecular investigation of its etiology. These findings motivate continued investigation into the functional roles of these proteins, charting a path toward molecularly informed treatment of dementia’s most distressing behavioral dimensions.

## Data Availability

Proteomics data used in this study are available in Synapse (https://www.synapse.org/) and can be located using Synapse ID syn73569657. A Synapse user account is required to access the data.

## Acknowledgements

We are grateful to the research volunteers and their families, who made this study possible. Figure 1 partially created in BioRender. Vattathil, S. (2026) https://BioRender.com/wddceyi

## Data Availability

Proteomics data used in this study are available in Synapse (https://www.synapse.org/) and can be located using Synapse ID <u>syn73569657</u>. A Synapse user account is required to access the data.

Clinical data is under restricted access to protect patient privacy. Data for each cohort can be requested as follows:

Layton Aging and Alzheimer’s Disease Research Center – using the form at https://www.ohsu.edu/alzheimers-disease-research-center/data-resources

Rush Alzheimer’s Disease Center (RADC) memory clinic – using the form at https://www.radc.rush.edu/requests/data.htm

Emory Goizuieta Alzheimer’s disease research center – using the form at https://alzheimers.emory.edu/research/for-researchers/data-request-form.html.

## Code Availability

Analyses were performed using R version 4.4.2 or other publicly available software and databases as noted. Code is available on request.

R packages: car (3.1-3), clusterProfiler (4.14.6), geepack (1.3.13), mixmeta (1.2.0), polycor (0.8-1), psych (2.5.3), qvalue (2.38.0), sva (3.54.0), WGCNA (1.73)

Other software: FragPipe (https://github.com/Nesvilab/FragPipe); METAL (https://csg.sph.umich.edu/abecasis/Metal/)

Databases: UniProt, downloaded February 2019 supplemented with APOE 2 and 4 and Abeta 40 and 42 isoforms

## Author Contributions

Conceptualization, Funding Acquisition, Supervision: APW, TSW; Resources: DAB, RSW, RLW; Data Collection, Data Curation: DMD, MG, NTS; Data curation, Formal analysis, Visualization: SMV; Writing – original draft: SMV, APW; Writing – review and editing: all authors.

## Additional Data Files

Additional Data File 1. Results from differential expression analysis for individual BPSD domains. Additional Data File 2. Results from differential expression analysis for individual BPSD domains in females only.

Additional Data File 3. Results from differential expression analysis for individual BPSD domains in males only.

Additional Data File 4. Results from differential expression analysis for individual BPSD domains adjusted for APOE ε4 expression.

Additional Data File 5. Results from differential expression analysis for BPSD latent factors.

